# The TGF-β1-oxidative stress axis underlies accelerated senescence of endothelial cells exposed to serum from hypertensive patients

**DOI:** 10.1101/2025.03.23.25324491

**Authors:** Paweł Uruski, Justyna Mikuła-Pietrasik, Andrzej Tykarski, Krzysztof Książek

## Abstract

**Aims:** There is a bidirectional link between hypertension (HT) and cellular senescence of endothelial cells (ECs). However, the mechanisms underlying EC senescence in patients with HT are not yet fully understood.

**Methods and Results:** We analyzed serum from 71 patients with primary HT and compared it to serum from 25 healthy donors to assess its effects on EC biology, including biomarkers, signaling pathways, and cellular senescence effectors. Our findings revealed that exposing ECs to serum from HT patients (20% for 72 h) impaired cell viability while enhancing proliferation, migration, and tubulogenesis. This effect is accompanied by increased expression of HIF-1α. Additionally, HT serum potentiated the expression of the senescence marker SA-β-Gal, shortened telomeres, and up-regulated cell-cycle inhibitors p16, p21, and p53. Regarding the signaling pathways, HT serum activated ERK1/2, p38 MAPK, AP-1/c-jun, and Notch1. Indices of oxidative stress in ECs treated with HT serum also increased, as indicated by elevated production of superoxides, activation of antioxidants (SOD, CAT), and accumulation of oxidized DNA, proteins, and lipids. Furthermore, mitochondria in these cells displayed decreased inner membrane potential and increased biogenesis, likely due to enhanced activity of PGC-1α. The activity of respiratory chain enzymes, including cytochrome c oxidase and NADH dehydrogenase, was also elevated. When HT serum-treated ECs were pre-incubated with the ROS scavenger PBN, the activity of SA-β-Gal decreased. A similar reduction in SA-β-Gal activity was observed when HT serum, which contained elevated levels of TGF-β1, was pre-incubated with a TGF-β1-neutralizing antibody. Importantly, exogenous TGF-β1, administered at a dose corresponding to its concentration in HT serum, induced senescence in ECs.

**Conclusions:** Our results indicate that serum from HT patients promotes senescence in ECs through mechanisms related to TGF-β1 and oxidative stress signaling.

## INTRODUCTION

Primary hypertension (HT) is a complex disease that can lead to serious health complications, including cardiovascular issues, chronic kidney disease, and, ultimately, death [1]. As blood pressure (BP) tends to rise progressively with age—due to various changes in artery structure and function [2]—the incidence of hypertension is particularly high among the elderly [3]. Several factors contribute to increasing BP, such as genetic predisposition, activation of the renin-angiotensin-aldosterone system (RAS), heightened sympathetic nervous system activity, and kidney filtration rate impairment. At the cellular level, HT is linked to dysfunction in vascular cells, especially endothelial cells (ECs). This dysfunction is driven by several underlying mechanisms, with inflammation and oxidative stress being the most significant factors [4, 5].

Recent evidence suggests that HT may be linked to cellular senescence of ECs [6]. Cellular senescence refers to a state where cells permanently exit the cell cycle due to DNA damage or stress, displaying distinct morphological, molecular, and biochemical signatures. Senescent cells typically accumulate with aging and are associated with the development of various age-related diseases [7]. Regarding HT, aging leads to a decline in endothelial-dependent dilation (EDD) [8]. This decline is thought to result from the accumulation of senescent ECs, which exhibit an inflammation-promoting secretory profile and reduced levels of vasodilators, such as endothelial nitric oxide synthase (eNOS), nitric oxide (NO), and prostacyclin. Additionally, factors such as oxidative stress and telomere uncapping contribute to this process [6, 9].

ECs play a dual role in HT pathogenesis, acting both as contributors to the disease and as its primary victims. This vulnerability primarily arises from mechanical cell damage induced by high BP, which can lead to various abnormalities [10], including cellular senescence [11]. Importantly, the effects of HT-induced EC dysfunction extend beyond the cardiovascular system. For example, recent studies have shown that ECs exposed to hypertensive serum undergo molecular and functional changes leading to increased cancer progression [12].

Taking all these facts into account, there is likely a bidirectional relationship between HT and the cellular senescence of ECs. However, the molecular mechanisms through which HT may induce senescence in ECs remain unclear. To investigate this, we designed an in vitro study to determine whether serum from HT individuals, which has a distinct biochemical composition compared to normal serum [13], could accelerate senescence in ECs. Our study aimed to examine the short-term effects (over 72 h) of HT serum on key functional properties of ECs, including viability, proliferation, migration, and tubulogenesis. We also evaluated biomarkers of cellular senescence, focusing on DNA damage response (DDR) elements, telomeres, and cell cycle inhibitors, alongside the activation of senescence-associated signaling pathways. Additionally, we assessed a wide range of parameters linked to oxidative stress and mitochondrial function. To further understand these processes, we conducted specific intervention tests to identify agents or phenomena whose targeting could potentially inhibit HT-related senescence in ECs.

## MATERIALS AND METHODS

### Chemicals and consumables

All chemicals and culture plastics were sourced from Sigma (St. Louis, MO), unless otherwise noted. The recombinant forms of human IL-6, TGF-β1, GRO-1, and HGF, along with a specific TGF-β1 neutralizing antibody, were obtained from R&D Systems (Abingdon, UK).

### Characteristics of HT patients and healthy volunteers

This study involved two groups: 25 healthy volunteers (control group) and 71 patients with newly diagnosed primary hypertension (HT group). Hypertension was diagnosed based on the 2018 Guidelines of the European Society of Cardiology [14], defined as systolic blood pressure (SBP) values of ≥ 140 mmHg and/or diastolic blood pressure (DBP) values of ≥ 90 mmHg. Exclusion criteria included: signs or a diagnosis of secondary hypertension, prior hypotensive treatment, chronic kidney disease with an estimated glomerular filtration rate (eGFR) < 60 ml/min/1.73m², individuals who were underage, pregnant, or breastfeeding. The institutional bioethics committee approved the study (consent number 415/17, Aug. 6, 2017), and all participants provided written informed consent. Comprehensive medical interviews and examinations were conducted for each patient, focusing on potential causes of secondary hypertension and current medications. Office blood pressure measurements were recorded using the Omron 705 IT device, taking the average of three consecutive readings while the participants were seated. Blood samples from the hypertensive patients were collected at the time of diagnosis, centrifuged immediately, and the serum was stored in aliquots at -80°C for future analysis. The characteristics of both the control and hypertensive groups are detailed in Table 1.

**Table I.**
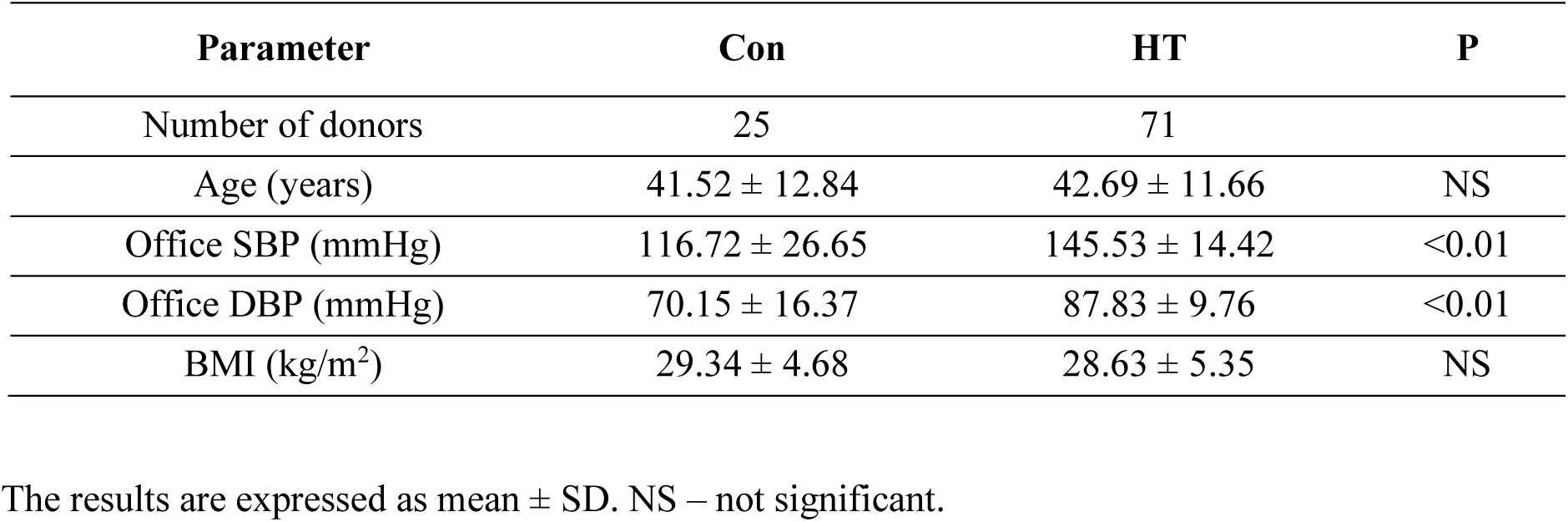
Basic clinical characteristics of HT patients and control volunteers.

| Parameter | Con | HT | P |
| --- | --- | --- | --- |
| Number of donors | 25 | 71 |  |
| Age (years) | 41.52 ± 12.84 | 42.69 ± 11.66 | NS |
| Office SBP (mmHg) | 116.72 ± 26.65 | 145.53 ± 14.42 | <0.01 |
| Office DBP (mmHg) | 70.15 ± 16.37 | 87.83 ± 9.76 | <0.01 |
| BMI (kg/m<sup>2</sup>) | 29.34 ± 4.68 | 28.63 ± 5.35 | NS |

The results are expressed as mean ± SD. NS – not significant.

### Endothelial cell (EC) culture and experimental protocol

Human umbilical vein endothelial cells (HUVECs) were purchased from the American Type Culture Collection (Rockville, MD, USA). The cells were cultured in EBM™-2 Basal Medium and EGM™-2 SingleQuots™ Supplements (Lonza, Walkersville, MD). During the experiments, young (early-passage) ECs that reached ∼80% confluency were exposed to the growth medium enriched with 20% serum from HT patients and healthy volunteers for 72 h, as described in [15].

### Cell death and apoptosis measurement

The incidence of apoptosis and necrosis in ECs treated with HT and Con sera was quantified using the Apoptotic/Necrotic/Healthy Cells Detection Kit (PromoKine; Heidelberg, Germany) under the Axio Vert.A1 fluorescent microscope (Carl-Zeiss, Jena, Germany).

### Examination of EC motility

The proliferation of ECs was assessed using the Cell Proliferation Kit I (PromoKine). To quantify their migration towards a chemotactic gradient generated by specific sera (20%), ChemoTx migration chambers (Neuro Probe, Gaithersburg, MD) were employed. In both analyses, fluorescence measurement was conducted using a SynergyTM 2 spectrofluorometer (BioTek Instruments, Winooski, VT, USA). Additionally, tube formation was evaluated using a Cultrex in vitro Angiogenesis Assay Tube Formation Kit (Trevigen, Gaithersburg, MD, USA). For detailed methodologies regarding EC proliferation, migration, and tubulogenesis measurements, refer to [16]. Furthermore, hypoxia-inducible factor-1 alpha (HIF-1) levels were quantified using a Human HIF-1 alpha ELISA kit (Biorbyt, Cambridge, UK), following the manufacturer’s instructions.

### Analysis of cellular senescence

The percentage of cells exhibiting cytosolic activity of senescence-associated β-galactosidase (SA-β-Gal) was quantified following the method outlined by Dimri et al. [17]. The enzyme’s activity was assessed using a fluorescence-based approach described by Gary and Kindell [18]. In certain experiments, we measured SA-β-Gal activity after exposing cells to exogenous, recombinant forms of human cytokines, including IL-6, TGF-β1, GRO-1, and HGF, at concentrations reflecting the average levels found in serum from HT patients. In a separate series of experiments, SA-β-Gal activity was re-evaluated in ECs that were subjected to serum from HT patients, which had been pre-incubated with a specific TGF-β1 neutralizing antibody (400 ng/ml) for 4 h [19]. Moreover, the enzyme’s activity was investigated in cells that were pre-treated with the reactive oxygen species (ROS) spin-trap scavenger N-tert-butyl-alpha-phenylnitrone (PBN, 800 µM) for 4 h [20].

We also evaluated key elements of the senescence-associated DNA damage response (DDR), specifically analyzing foci of the phosphorylated variant of histone H2A.X (γ-H2A.X) and p53-binding protein 1 (53BP1). This was performed using anti-γ-H2A.X (cat #ab2893, Abcam, Cambridge, UK) and anti-53BP1 antibodies (cat #NB100-304, Novus Biologicals, Abingdon, UK), as described in [21]. Telomere length was assessed using the Absolute Human Telomere Length Quantification qPCR Assay Kit (ScienCell, Carlsbad, CA). Genomic DNA was extracted using the Genomic Mini Kit (A&A Biotechnology, Gdynia, Poland), and the PCR reaction was executed with a LightCycler 96 (Roche, Penzberg, Germany), utilizing FastStart Essential DNA Green Master (Roche) and 5 ng of DNA. The Comparative Quantification Cycle Value method was employed to determine telomere length. Finally, the expression of cell cycle inhibitors p16, p21, and p53 was detected through immunofluorescence in cells incubated with specific antibodies: rabbit anti-human p16 (#ab108349; Abcam, Cambridge, UK), p21 (#2947, Cell Signaling), and p53 (#2527, Cell Signaling Technology, Danvers, MA), following the protocols described in [22, 23].

### Quantification of oxidative stress-related parameters

The study investigated the generation of mitochondrial superoxides and cellular peroxides, as well as mitochondrial mass and inner membrane potential (ΔΨm). This was achieved by measuring fluorescence generated by various indicators: MitoSox Red, dihydrorhodamine 123 (DHR), 10-n-nonyl-acridine orange (NAO), and 5,5′,6,6′-tetrachloro-1,1′,3,3′-tetraethylbenzimidazolylcarbocyanine iodide (JC-1). Detailed protocols for these analyses can be found in reference [24]. The concentration of 8-hydroxy-2’-deoxyguanosine (8-OH-dG), a significant product of DNA oxidation, was quantified using the DNA Damage (8OHdG) ELISA Kit from Biorbyt Ltd. (Cambridge, UK). To evaluate oxidative modifications of cellular lipids, 8-isoprostane (8-iso Prostaglandin F2α) was quantified using the 8-Isoprostane ELISA kit from Cayman Chemical (Ann Arbor, MI, USA). The content of carbonylated proteins was determined with a dedicated assay from Cayman Chemical as well. Additionally, the enzymes involved in the formation of mitochondrial reactive oxygen species (ROS) — such as cytochrome c oxidase and NADH dehydrogenase — as well as those responsible for mitochondrial biogenesis, specifically peroxisome proliferator-activated receptor gamma coactivator-1 alpha (PGC-1α), were quantified using kits from Wuhan EIAab Science Co., Ltd. (Wuhan, China). Finally, enzymes that play a role in antioxidative reactions, namely superoxide dismutase (SOD) and catalase (CAT), were measured using assays from Cayman Chemical. All commercial assays were performed following the manufacturer’s instructions.

### Immunoassays

The concentration of intercellular adhesion molecule 1 (ICAM-1), vascular cell adhesion molecule 1 (VCAM-1), E-selectin, P-selectin, plasminogen activator inhibitor 1 (PAI-1), endothelin-1 (ET-1), growth-related oncogene 1 (GRO-1), transforming growth factor β1 (TGF-β1), tumor necrosis factor α (TNF-α), interleukin 6 (IL-6), interleukin 8 (IL-8), hepatocyte growth factor (HGF), platelet-derived growth factor (PDGF), basic fibroblast growth factor (bFGF), epidermal growth factor (EGF), insulin-like growth factor 1 (IGF-1), angiopoietin 1, monocyte chemoattractant protein-1 (MCP-1), and vascular endothelial growth factor (VEGF) in serum was measured using appropriate DuoSet^®^ Immunoassay Development kits (R&D Systems), according to manufacturer’s instructions.

### Analysis of senescence-associated kinases and transcription factors

The activation of AKT, AP-1/c-jun, ERK1/2, p38 MAPK, NF-κB/p65, Notch1, and STAT3 was quantified using total/phospho InstantOne ELISA^TM^ kits from Invitrogen (Eugene, OR, USA). JAK1 activation was quantified using a respective kit from Abcam (Cambridge, UK).

### Statistics

Statistical analysis was conducted using GraphPad Prism version 9.5.1 software (GraphPad Software, San Diego, USA). The Kolmogorov-Smirnov test was used to assess the normality of continuous data distributions. If the distribution was normal, we applied the unpaired t-test or one-way ANOVA. In cases where at least one group exhibited a non-Gaussian distribution, we utilized non-parametric tests, specifically the Mann-Whitney test or the Kruskal-Wallis test. The Wilcoxon signed-rank test was employed as appropriate. Due to the unequal number of samples in the HT and Con groups, a power analysis was performed to confirm that sample sizes were sufficient to detect significant differences. Results are presented as means ± standard deviation (SD), with differences deemed statistically significant at a P-value of less than 0.05.

## RESULTS

### Serum from HT patients impairs viability and promotes motility of ECs

ECs were exposed to control (Con) serum and hypertensive (HT) serum for 72 hours, after which we quantified the incidence of apoptotic and necrotic cell death. Notably, both types of cell death were assessed in a mixture of adherent cells and those that had detached from the dish and were floating in the growth medium. Measurements utilizing annexin V for apoptosis and ethidium homodimer III for necrosis demonstrated that HT serum induces both forms of cell death in approximately 20% of the cells (Fig. 1A).

**Figure 1.**
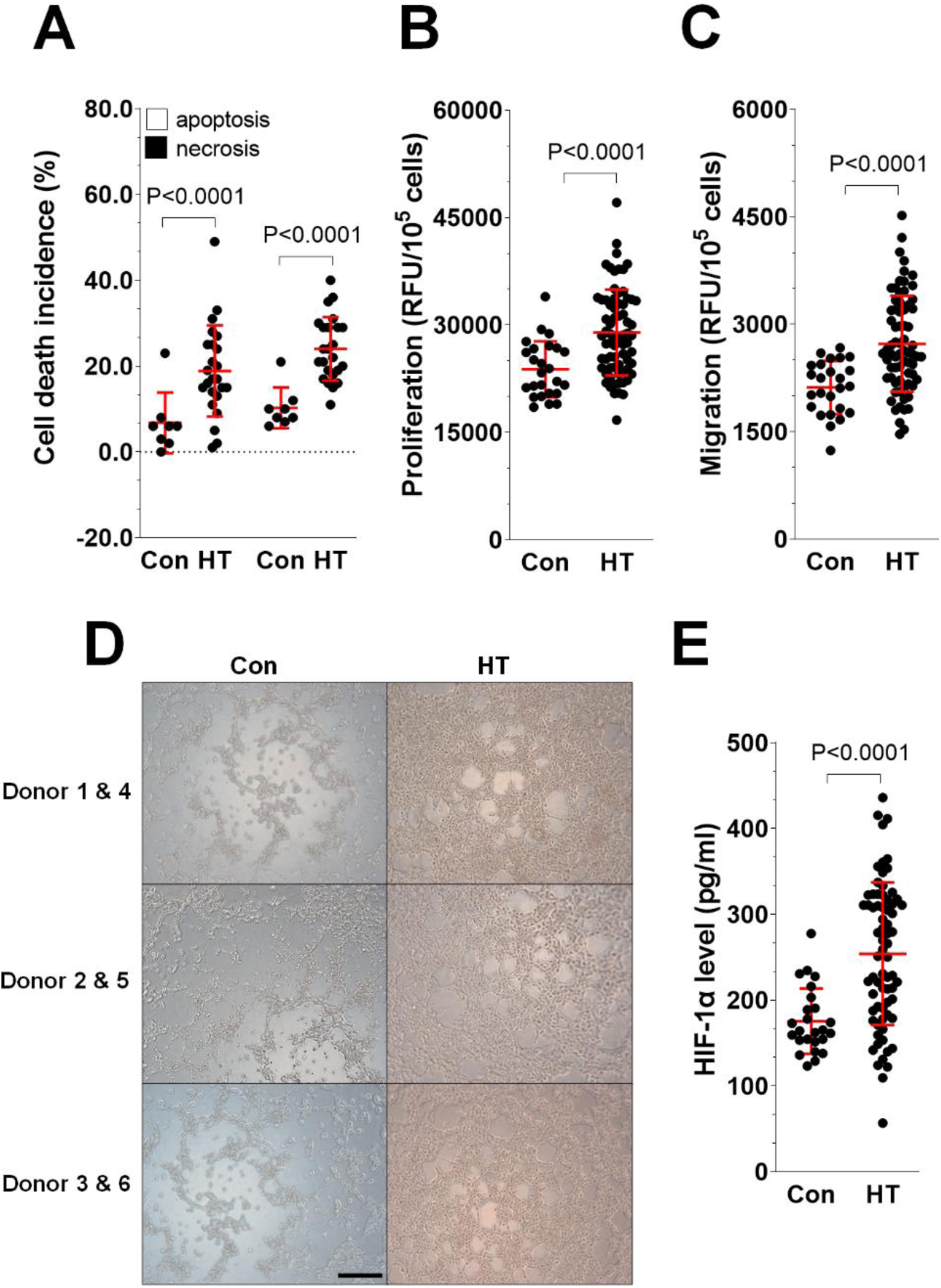
Functional properties of ECs subjected to serum from hypertensive (HT) and control (Con) donors. ECs were exposed to 20% hypertensive (HT) serum and control (Con) serum for 72 h. Following this exposure, we assessed several outcomes: the incidence of apoptosis and necrosis (A), proliferation (B), migration (C), and the ability to form microtubules (D). Additionally, we quantified the expression of HIF-1α (E). The experiments utilized sera from 8 Con donors and 24 HT donors for part A; from 25 Con donors and 71 HT donors for part B, C, and E; and from 6 Con donors and 6 HT donors for part D. Scatter dot plots include mean ± SD values. Magnification × 100. Scale bar, 200 μm. RFU – Relative Fluorescence Units.

Subsequently, we evaluated the motility of ECs, focusing on their proliferation and migration. A comparative analysis revealed that HT serum significantly enhances both behaviors of the endothelial cells (Fig. 1B, C). This stimulatory effect was associated with an increased ability of cells exposed to HT serum to form microtubules (Fig. 1D) and a higher expression level of the transcription factor HIF-1α (Fig. 1E).

### Serum from HT patients promotes cellular senescence in ECs

Cellular senescence is a response of cells to various internal and external stressors [25]. ECs exposed to HT serum exhibited several markers of senescence. Notably, there was an increase in the percentage of SA-β-Gal-positive cells (Fig. 2A, C) and elevated activity of the enzyme itself (Fig. 2B). Additionally, these cells showed shortened telomeres (Fig. 2D) and activation of key cell cycle inhibitors, including p16, p21, and p53 (Fig. 2G, H, I, J). Importantly, HT serum exposure did not trigger a DNA damage response in the ECs, as indicated by unchanged levels of histone γ-H2A.X and 53BP1 (Fig. 2E, F).

**Figure 2.**
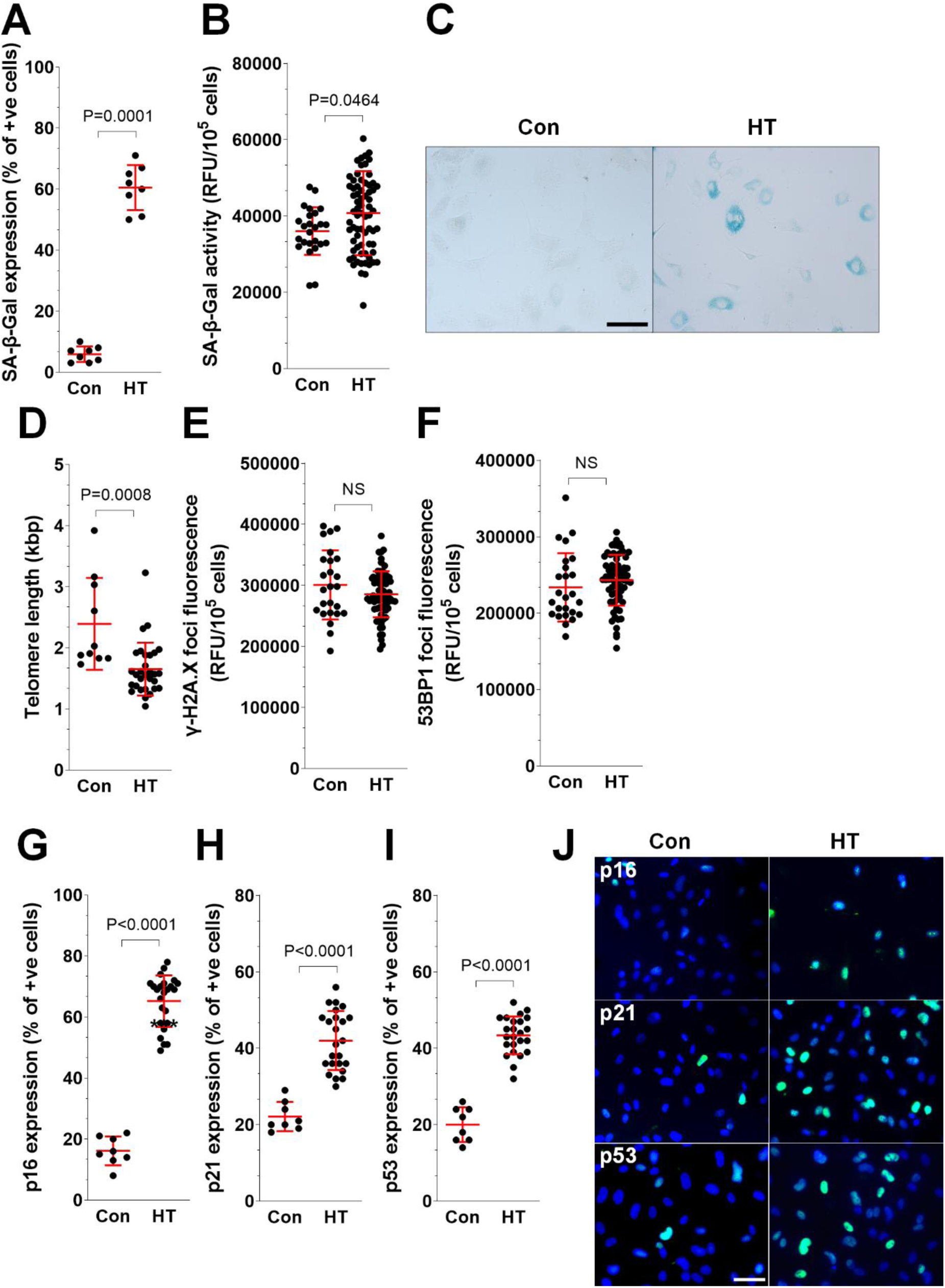
Effect of hypertensive (HT) and control (Con) serum on EC senescence. ECs were exposed to 20% serum from healthy controls (Con) and patients with hypertension (HT) for 72 h. We then evaluated various parameters: the expression (A) and activity (B) of SA-β-Gal, telomere length (D), the expression of histone H2A.X (E) and 53BP1 (F), and the percentage of cells expressing p16 (G), p21 (H), and p53 (I). Representative images of SA-β-Gal staining (C) and p16, p21, p53 staining (J) were also captured. The experiments were conducted using serum samples from 8 Con and 8 HT donors for measurements A; 25 Con and 71 HT donors for measurements B, E, and F; 10 Con and 30 HT donors for measurement D; and 8 Con and 24 HT donors for measurements G, H, and I. Furthermore, the micrographs shown in panels C and J were taken from experiments using sera from 6 Con and 6 HT donors. Scatter dot plots include mean ± SD values. Magnification × 200 (C) and × 400 (J). Scale bar, 200 (C) and 500 (J) μm. RFU – Relative Fluorescence Units. NS – not significant.

### Serum from HT patients induces senescence-associated signaling pathways

To investigate the signaling pathways involved in the cellular senescence of ECs triggered by HT serum, we quantified the activation of several kinases and transcription factors associated with senescence. Our experiments showed that HT serum activates ERK1/2 and p38 MAPK kinases, as well as the transcription factors AP-1/c-jun and Notch1. In contrast, the activity of AKT, JAK1, NF-κB/p65, and STAT3 remained unchanged (Fig 3).

**Figure 3.**
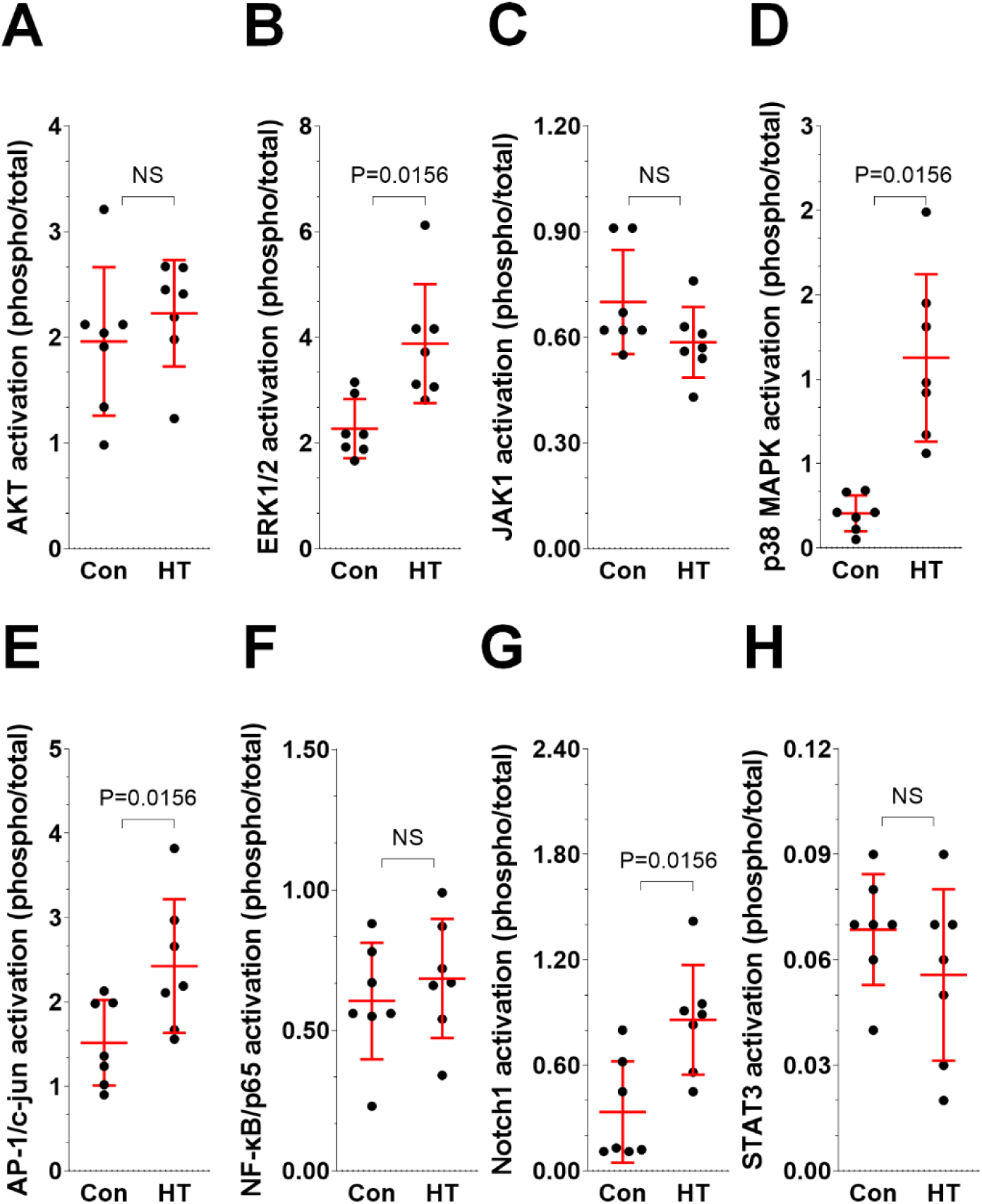
Effect of hypertensive (HT) and control (Con) serum on senescence-associated signaling pathways in ECs. ECs were exposed to 20% hypertensive (HT) serum and control (Con) serum for 72 h. Following this exposure, the activation levels of several signaling pathways were quantified, including AKT (A), ERK1/2 (B), JAK1 (C), p38 MAPK (D), AP-1/c-jun (E), NF-κB/p65 (F), Notch1 (G), and STAT3 (H). The experiments utilized serum samples from 7 donors each for the Con and HT groups. Scatter dot plots include mean ± SD values. NS – not significant.

### Serum from HT patients induces mitochondrial oxidative stress in ECs

Oxidative stress is a key factor in the acceleration of cellular senescence [26]. ECs exposed to HT serum showed an increased production of mitochondrial superoxides, as indicated by enhanced MitoSOX red fluorescence. In contrast, the levels of cellular peroxides, measured by DHR fluorescence, remained unchanged. To counteract the elevated reactive oxygen species (ROS), the activities of antioxidative enzymes, specifically superoxide dismutase (SOD) and catalase (CAT), were up-regulated, likely as a compensatory mechanism. Further evidence of heightened oxidative stress in HT serum-treated ECs includes significant oxidative modifications to cellular macromolecules. These cells displayed increased levels of oxidative damage: DNA showed signs of damage through 8-hydroxydeoxyguanosine (8-OH-dG), while proteins exhibited carbonylation, and lipids demonstrated increased levels of 8-isoprostane (Table II).

**Table II.** The effect of HT serum on oxidative stress-related parameters in cultured ECs.

| Parameter | Con | HT | P |
| --- | --- | --- | --- |
| 8-isoprostane (pg/ml) | 18.57 ± 8.65 | 35.49 ± 7.47 | <0.001 |
| 8-OH-dG (ng/ml) | 21.23 ± 3.11 | 29.04 ± 3.52 | <0.001 |
| Carbonylated proteins (nmol/ml) | 76.55 ± 66.26 | 169.90 ± 187.43 | 0.008 |
| CAT (nmol/min/ml) | 12.66 ± 8.76 | 33.12 ± 16.96 | <0.001 |
| Cytochrome c oxidase (ng/ml) | 7.39 ± 3.51 | 16.31 ± 4.21 | <0.001 |
| DHR (RFU/10<sup>5</sup> cells) | 267470 ± 63435 | 266747 ± 38394 | NS |
| JC-1 (RFU/10<sup>5</sup> cells) | 3.82 ± 0.99 | 2.54 ± 0.89 | <0.001 |
| MitoSOX red (RFU/10<sup>5</sup> cells) | 409329 ± 64410 | 372642 ± 57689 | 0.009 |
| NADH dehydrogenase (ng/ml) | 5.99 ± 2.35 | 8.81 ± 5.17 | 0.003 |
| NAO (RFU/10<sup>5</sup> cells) | 25994 ± 3641 | 28476 ± 4211 | 0.0027 |
| PGC-1α (ng/ml) | 7.916 ± 1.24 | 10.77 ± 3.39 | <0.001 |
| SOD (U/ml) | 0.83 ± 0.33 | 1.15 ± 0.20 | <0.001 |

The experiments were performed using sera from HT patients (n=71) and control donors (n=25). The results are expressed as mean ± SD. NS – not significant.

The oxidative stress induced by HT serum in ECs appears to originate from mitochondrial dysfunction. These cells showed heightened activity of NADH dehydrogenase and cytochrome c oxidase, both of which contribute to mitochondrial ROS production. Dysfunctional mitochondria in these cells produced less ATP, indicated by decreased values of inner membrane potential (ΔΨm), as seen through JC-1 fluorescence. To compensate for this decreased ATP production, the cells increased the biogenesis of new mitochondria, as evidenced by enhanced NAO fluorescence and the elevated enzymatic activity of PGC-1α (Table II).

### Serum from HT patients displays pro-inflammatory characteristics

Several parameters related to local and systemic inflammation were measured in the HT serum and compared to those in healthy donors. The serum from HT patients exhibited a distinctly pro-inflammatory profile, characterized by elevated levels of several markers. Specifically, there were increased values of C-reactive protein (CRP), as well as higher concentrations of various cytokines (such as IL-6), chemokines (including IL-8, GRO-1, and MCP-1), growth factors (such as HGF, IGF-1, PDGF, and TGF-β1), and adhesion molecules (including ICAM-1, VCAM-1, E-selectin, and P-selectin) (Table III).

**Table III.** The effect of HT serum on the secretory profile of cultured ECs.

| Parameter | Con | HT | P |
| --- | --- | --- | --- |
| Angiopoietin 1 (pg/ml) | 7663 ± 1858 | 8787 ± 1949 | 0.002 |
| CRP | 1.54 ± 1.47 | 3.68 ± 3.61 | 0.0084 |
| EGF (pg/ml) | 191 ± 46.6 | 218 ± 54.7 | 0.02 |
| Endothelin-1 (pg/ml) | 19 ± 18 | 15 ± 4 | NS |
| E-selectin (pg/ml) | 2092 ± 470 | 4125 ± 469 | <0.001 |
| bFGF (pg/ml) | 66 ± 12 | 75 ± 21 | NS |
| GRO-1 (pg/ml) | 382 ± 199 | 505 ± 276 | 0.03 |
| HGF (pg/ml) | 405 ± 303 | 648 ± 173 | <0.001 |
| sICAM-1 (pg/ml) | 2252 ± 601 | 2638 ± 377 | <0.001 |
| IGF-1 (pg/ml) | 702 ± 560 | 1144 ± 260 | <0.001 |
| IL-6 (pg/ml) | 94 ± 23 | 107 ± 21 | 0.007 |
| IL-8 (pg/ml) | 206 ± 52 | 271 ± 106 | <0.001 |
| MCP-1 (pg/ml) | 909 ± 413 | 1073 ± 438 | NS |
| P- selectin (pg/ml) | 2515 ± 433 | 3269 ± 943 | <0.001 |
| PAI-1 (pg/ml) | 8719 ± 2287 | 9007 ± 2776 | NS |
| PDGF (pg/ml) | 96 ± 19 | 133 ± 48 | <0.001 |
| TGF-β1 (pg/ml) | 764 ± 166 | 1142 ± 180 | <0.001 |
| TNF-α (pg/ml) | 1.57 ± 0.66 | 1.56 ± 0.65 | NS |
| VCAM-1 (pg/ml) | 273 ± 48 | 340 ± 53 | <0.001 |
| VEGF (pg/ml) | 156 ± 82 | 208 ± 112 | 0.01 |

The measurements were performed using sera from HT patients (n=71) and control donors (n=25).
The results are expressed as mean ± SD. NS – not significant.

Additionally, the HT serum displayed higher concentrations of pro-angiogenic agents such as angiopoietin 1, epidermal growth factor (EGF), and vascular endothelial growth factor (VEGF). In contrast, the levels of endothelin-1, basic fibroblast growth factor (bFGF), MCP-1, plasminogen activator inhibitor-1 (PAI-1), and tumor necrosis factor-alpha (TNF-α) were similar in both groups (Table III).

### HT-related acceleration of ECs’ senescence is triggered by TGF-β1 and oxidative stress

Four cytokines that were found in increased concentrations in HT serum compared to Con serum have been previously identified as potential inducers of cellular senescence in various cell types. These cytokines are IL-6 [27], TGF-β1 [28], GRO-1 [29], and HGF [30]. To investigate whether these factors contribute to senescence induction in ECs exposed to HT serum, recombinant forms of these cytokines were administered at concentrations corresponding to those found in serum samples. The ECs were treated for 72 h, after which the activity of the senescence biomarker SA-β-Gal was assessed.

The results indicated that IL-6, GRO-1, and HGF did not induce senescence in ECs (data not shown). However, recombinant TGF-β1 significantly increased SA-β-Gal activity (Fig. 4A). To further confirm whether TGF-β1 from HT serum was responsible for the observed senescence in ECs, intervention tests were performed using specific neutralizing antibodies against TGF-β1 added to the serum. This approach demonstrated that neutralization of TGF-β1 in HT serum reduced SA-β-Gal activity to levels comparable to the control group (Fig. 4B).

**Figure 4.**
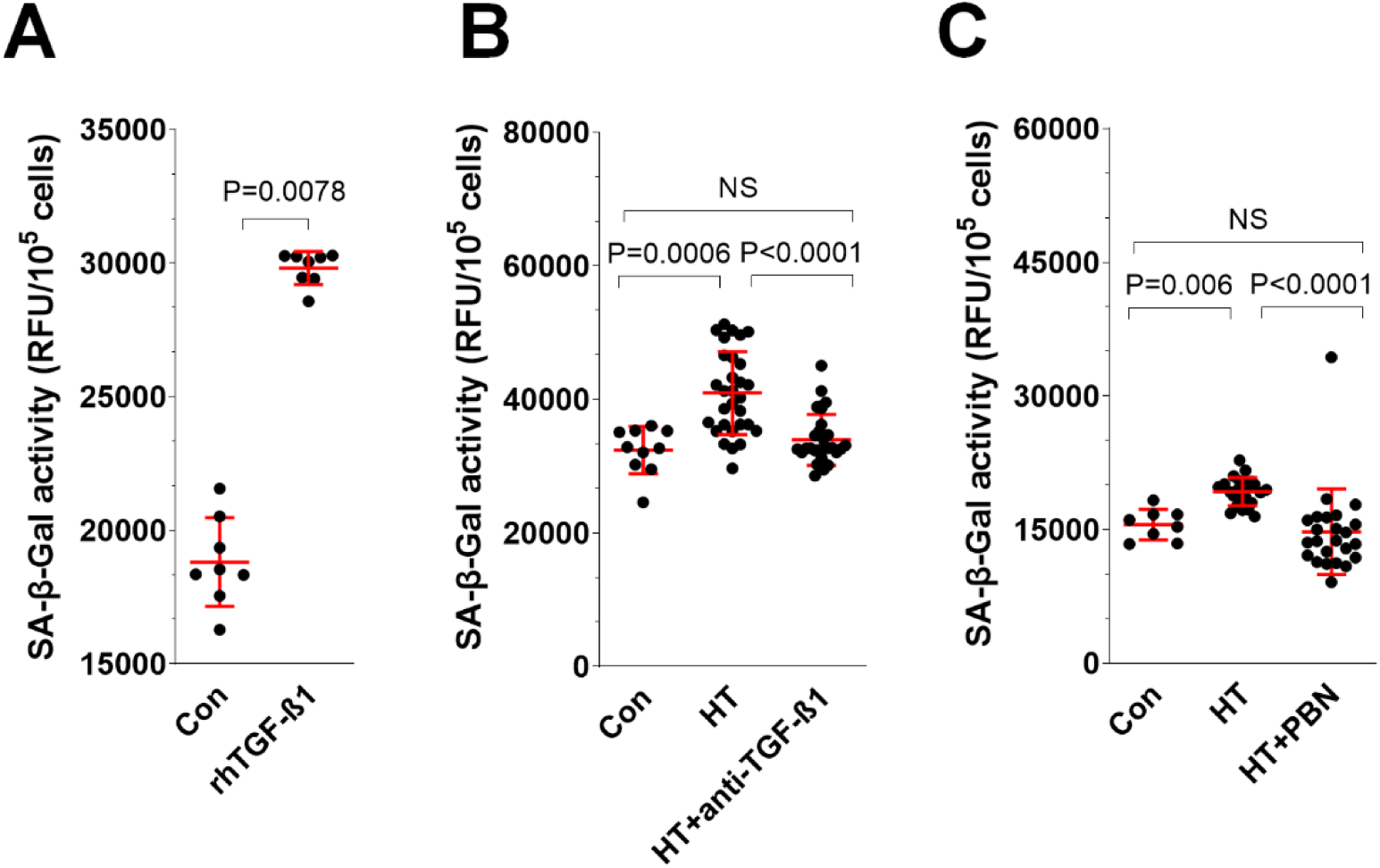
The role of TGF-β1 and oxidative stress in HT serum-induced senescence of ECs. The activity of SA-β-Gal in ECs exposed to recombinant TGF-β1 was assessed (A). Additionally, the activity of SA-β-Gal was measured in ECs exposed to Con serum, HT serum, and HT serum that had been pre-incubated with a specific anti-TGF-β1 neutralizing antibody (B). Furthermore, the activity of SA-β-Gal was evaluated in ECs exposed to Con and HT serum, as well as in cells treated with HT serum after pre-incubation with the ROS scavenger, PBN (C). The experiments included sera from 8 Con and 8 HT donors for part A; 10 Con, 30 HT, and 30 HT + anti-TGF-β1 for part B; and 8 Con, 24 HT, and 24 HT + PBN donors for part C. Scatter dot plots include mean ± SD values. RFU – Relative Fluorescence Units, NS – not significant.

Additionally, since HT serum was found to enhance mitochondrial oxidative stress in ECs, further intervention tests were conducted with a spin-trap ROS scavenger, PBN. ECs were pretreated with PBN before exposure to HT serum. These experiments showed that the elevated SA-β-Gal activity in response to HT serum declined to levels typical of Con serum when cells were protected against oxidative stress with PBN (Fig. 4C).

## DISCUSSION

The relationship between hypertension (HT) and cellular senescence in endothelial cells (ECs) is widely believed to be bidirectional [6]; however, the specific molecular mechanisms through which HT promotes senescence remain unclear. In this study, we employed a well-established protocol using in vitro endothelial cell (HUVEC) exposure to human serum [12, 31, 32] to investigate how serum from HT patients modifies the basic activity of ECs and whether it induces senescence in these cells. Our experiments revealed that serum from HT patients is detrimental to ECs, as it increased cell death through mechanisms such as apoptosis and necrosis. This heightened mortality was countered by increased proliferation and migration of the ECs, which also enhanced their ability to form microtubules. Increased expression of the transcription factor HIF-1α is a plausible driver of these effects [33]. However, while enhanced cell proliferation may initially seem beneficial, it leads to a more rapid depletion of the cells’ finite capacity for division, suggesting that these compensatory reactions could contribute to premature senescence. This phenomenon of short-term heightened proliferation followed by premature exhaustion has parallels in other cell types; for instance, mesangial cells exposed to harmful high glucose conditions showed a similar pattern of proliferation [34]. Likewise, peritoneal mesothelial cells also experienced increased divisions when exposed to high glucose, ultimately leading to a premature loss of replicative capacity and senescence [35, 36].

In line with this assumption, ECs exposed to HT serum exhibited accelerated senescence. This process was evident through an increase in the senescence biomarker, SA-β-Gal, and primarily occurred in a telomere-dependent manner, as demonstrated by telomere erosion and the activation of the p53-p21 effectory axis. This mechanism of senescence is characteristic of ECs [37], including those found in atherosclerotic lesions [38]. Interestingly, while senescence was induced by HT serum, it did not trigger the activation of the DNA damage response (DDR), which typically follows telomeric DNA disruption [39]. We speculate that the absence of DDR activation may be due to the relatively short duration of the experiment.

The activation of EC senescence is associated with the induction of specific signaling molecules, including members of the MAP kinase family, such as ERK1/2 and p38, as well as the transcription factors AP-1/c-jun and Notch1. Notably, the activation of ERK1/2 signaling may also contribute to the increased motility of ECs [40]. Meanwhile, p38 MAPK, a key kinase involved in the development of the senescence-associated secretory phenotype (SASP) [41], may play a role in the secretory characteristics of ECs. This secretory phenotype can facilitate the previously observed pro-cancerous changes in these cells when treated with HT serum [12].

Oxidative stress is recognized as a key factor in telomere-dependent cellular senescence [26, 42] and is closely linked to the pathophysiology of HT [43]. Consequently, our research focused on various parameters associated with this phenomenon. As anticipated, serum from HT patients induced oxidative stress in ECs, which was evidenced by increased production of ROS, such as superoxides, along with the accumulation of oxidized DNA, proteins, and lipids. This oxidative stress prompted a compensatory rise in the activities of two main antioxidant enzymes, superoxide dismutase (SOD) and catalase (CAT).

Mitochondria emerged as the primary source of this oxidative stress, as the elevated levels of superoxides likely resulted from mitochondrial dysfunction and the activation of a retrograde signaling response [44]. In cells treated with HT serum, mitochondria displayed decreased mitochondrial membrane potential (ΔΨm), indicating a reduced capacity to generate ATP. This de-energization triggered a compensatory stimulation of mitochondrial biogenesis mediated by PGC-1α [45], ultimately leading to increased production of ROS driven by increased activities of cytochrome c oxidase and NADH dehydrogenase [46]. The role of oxidative stress in the senescence of ECs induced by HT serum was further confirmed through intervention tests, which showed a decrease in SA-β-Gal activity when cells were treated with the ROS scavenger PBN.

The critical question at this stage is what triggers oxidative stress-dependent EC senescence in the composition of HT serum. Previous research has indicated that HT serum has a distinct biochemical profile compared to serum from healthy individuals, including differences in its cytokine composition [13]. Our findings corroborate earlier data, particularly concerning elevated levels of IL-6 [13] and TGF-β1 [47], while also broadening the range of cytokines and growth factors that are overproduced in HT serum. Table III lists the cytokines, chemokines, and growth factors present in higher concentrations in HT serum than in control serum. These factors are associated with processes such as angiogenesis (e.g., angiopoietin 1, VEGF) and inflammatory responses (e.g., sICAM-1, E-selectin). Among these, we selected four factors known to potentially induce cellular senescence: IL-6 [27], TGF-β1 [28], GRO-1 [29], and HGF [30]. However, intervention studies using recombinant forms of these proteins demonstrated that only TGF-β1 in HT serum is present at concentrations sufficient to induce senescence in ECs. Further experiments supported the idea that TGF-β1 acts as a likely trigger for EC senescence from HT serum. When this cytokine was neutralized in the serum with specific antibodies, there was a significant reduction in the biomarker of senescence, bringing it to levels typically seen in cells exposed to serum from healthy individuals. Importantly, there appears to be a plausible relationship between serum TGF-β1 levels and the oxidative stress-related signaling triggered in ECs treated with HT serum. Other studies have shown that TGF-β1 can induce ROS production and activate the p38 MAPK pathway [28]. This connection between TGF-β1 activity and increased oxidative stress in ECs has also been documented [48].

Overall, our study demonstrates that HT serum induces EC senescence in vitro. Notably, the experiments involved a relatively short exposure of 72 h to only 20% HT serum. It is reasonable to hypothesize that the pro-senescence effects could be significantly more pronounced in vivo with continuous exposure to 100% serum. Given the critical role of HT in the development of various cardiovascular conditions, such as atherosclerosis, myocardial infarction, and stroke, it is essential to investigate whether appropriate antihypertensive therapies could prevent or mitigate the impact of HT on EC senescence and its related co-morbidities. In this context, it is important to focus on the key factors identified in our study, specifically TGF-β1 and mitochondria-related oxidative stress.

## Data Availability

All data contained in the manuscript are available from the corresponding author.

## Acknowledgments

This research was funded by the National Science Centre, Poland, grant number 2018/02/X/NZ5/00403.

